# Defining Stimulability Testing in Voice Disorders: A Scoping Review

**DOI:** 10.1101/2025.10.23.25338650

**Authors:** Elizabeth D. Young

## Abstract

**Purpose:** Voice-specialized speech-language pathologists have been performing stimulability testing since at least the 1970s. However, definitions of stimulability testing vary widely within the current literature, leading to wide variability in stimulability testing practices. To determine the degree of consensus regarding stimulability testing, the current scoping review collected and analyzed the definitions of stimulability testing and stimulability testing outcomes amongst current peer-reviewed literature regarding voice disorders.

**Methods:** A literature search was conducted in PubMed, EBSCOhost, and Google Scholar for peer-reviewed articles containing the terms “stimulability” or “stimulable”. Articles including information regarding the behavioral assessment of patients with voice disorders were included.

**Results:** 88 articles were included in the review. Only 20% of articles provided a definition of stimulability testing or stimulability testing outcomes; over half (52%) mentioned search terminology in passing without any further definition or description, and the remaining quarter (27%) described stimulability testing or stimulability testing outcomes rather than providing a definition. Thematic analysis of stimulability testing suggested that most authors describe the practice as involving modifying patient behaviors, assessing patient responses, and using facilitators. While definitions of stimulability testing outcomes shared thematic similarities, details within the definitions varied substantially.

**Conclusion:** Stimulability testing and stimulability testing outcomes are infrequently defined within the current literature. Results of the thematic analysis suggest consensus on the common elements of stimulability testing, but not stimulability testing outcomes. Given the clinical importance of stimulability testing, increased care is needed in future work to provide full and adequate definitions of both stimulability testing and stimulability testing outcomes in future work.

## Introduction

The first use of the term “stimulability” is generally attributed to Milisen (1954)^1^ within the context of assessment for children with speech sound disorders. In the early 1970s, stimulability testing began to be adopted into other areas of speech-language pathology, particularly voice disorders. Many consider Daniel Boone’s “voice therapy facilitating techniques”, first introduced in his 1971 textbook^2^ as the basis of stimulability testing in voice disorders. Recent surveys of practicing voice-specialized speech-language pathologists (SLPs) found that 93% of respondents completed stimulability testing “always” or “almost always” with their patients, and 97% of respondents found it “important” or “very important” to their practice.^3^ Similarly, 94% of surveyed laryngologists report stimulability testing is performed “Always” or “Sometimes” in their practice, and 100% report there is a benefit to using it when working with patients with voice disorders.^4^ Thus, what began as a practice in speech-sound disorders has become extremely prevalent and highly valued for healthcare professionals involved in the care of the voice.

Despite the prevalence of stimulability testing in many areas of speech-language pathology, definitions of stimulability testing vary within the existing literature. For instance, Boone^2^ described stimulability testing as “probing continually” to find that one voice which sounds good”,^(p109)^ suggesting a potentially lengthy evaluation, whereas McDowell and colleagues^5^ define it as the assessment of a person’s ability to make “immediate changes to their voice”,^(p1)^ suggesting a much more rapid process. Both authors noted above specify voice as the behavioral target of stimulability testing; however, a wide variety of behavioral targets have been specified within stimulability testing definitions, including very broad targets (“…the assessment of an individual’s ability to modify a behavior when provided with models or cues“^6(p3)^) and very specific targets (“determine if the individual is capable of producing a louder voice easily and without straining”^7(p82)^). Some definitions also reference specific cueing approaches or contexts (“the desired behavior is elicited, then shaped, stabilized, and habituated using a hierarchical pattern”^8(p242)^) while others do not (e.g., “various facilitating techniques are introduced on a trial basis”^9(pxlix)^). Thus, many details about stimulability testing-–including factors such as expected time frame, behaviors to be evaluated, the context of evaluation, and the degree and types of therapeutic supports provided – are not standardized within the literature.

Because stimulability testing is not clearly defined, SLPs vary widely in how stimulability testing is performed. Survey-based research indicates that less than 10% of SLPs and less than 15% of laryngologists use a standardized protocol for stimulability testing in their current workplace.^3,4^ Further, in a qualitative study examining the stimulability testing practices of eight voice-specialized SLPs, three distinct approaches to stimulability testing were noted.^10^ This variability has led to poor conceptualization of the underlying theoretical constructs that stimulability testing is meant to measure. Stimulability testing outcomes have been described in a myriad of ways in the literature. Some examples of outcomes measured following stimulability testing include change in clinician-perceived voice quality following a facilitating technique,^11^ whether changes occurred in patient-perceived voice quality or voice feel,^12^ the degree of cueing required to elicit change in a patient’s voice,^5^ and, frequently, a combination of many factors.^5,13,14^ Thus, authors have described multiple theoretical constructs measured during stimulability testing (e.g., degree of perceptual change, patient responsiveness to cueing), often collapsed into a single outcome measure (e.g., “stimulable”). This reduces the clarity and precision with which the term “stimulable” can be used to communicate stimulability testing outcomes in clinical and research settings.

In some ways, a lack of standardization in stimulability testing practices is useful and necessary. Stimulability testing can be considered a form of dynamic assessment,^15^ where assessment decisions are made are based on previous patient responses. Framed this way, stimulability testing is most effective when not rigidly specified. However, because stimulability testing definitions vary so broadly within the literature, it is currently unclear what clinical practices constitute stimulability testing and what may constitute another practice entirely. For instance, an extended stimulability testing session may have no practical difference from a short voice therapy session. Additionally, without an understanding of the theoretical constructs commonly measured during stimulability testing, the active ingredients of stimulability testing cannot be systematically examined. For example, it is possible that responsiveness to cueing and the degree of perceptual voice change achieved during stimulability testing differentially affect patient responses to behavioral therapy. Thus, there is a need to evaluate the ex sting literature regarding stimulability testing to determine (a) the degree of consensus between definitions for both stimulability testing and stimulability testing outcomes, and (b) what specific elements of each definition have the highest degree of consensus. This will lay the groundwork for the standardization of the definition of stimulability testing and deepen our understanding of common stimulability testing outcomes.

The lack of uniformity used to describe stimulability testing within the literature has direct impact on clinical practice. Speech-language pathologists have long discussed stimulability testing as an invaluable tool for a variety of clinical functions, ranging from diagnostic (e.g., differential diagnosis^3,4,16,17^), to therapeutic (e.g., informing patient candidacy and prognosis for behavioral therapy ^4,12,18^) to meta-therapeutic (e.g., assessing patient learning methods, evaluating and improving patient motivation to attend behavioral therapy^4–6,12,13,19–21^). However, each of these proposed functions has limited substantiating evidence. For example, the relationship between stimulability testing and patient prognosis with behavioral voice therapy has only been examined by three studies to date.^5,12,22^ The results of these studies are mixed, with one suggesting a strong relationship,^22^ one trending towards a relationship,^12^ and one finding no relationship.^5^ One potential reason for these inconsistent findings is the poor standardization of what constitutes stimulability testing and stimulability testing outcomes. Each of the three studies used drastically different stimulability testing protocols and quantified the results in different ways. Thus, research investigating the valuable relationships between stimulability testing and its proposed clinical functions is limited by inconsistency in stimulability testing methods within the literature.

### The Current Study

Survey-based research indicates that both voice-specialized speech-language pathologists and laryngologists around the world perform stimulability testing frequently and value it as a clinical assessment tool.^3,4,23^ However, research surrounding stimulability testing is currently limited by the lack of standardization in both research and clinical practice. Authors have recently begun advocating for changes to stimulability testing practices, including standardization, in order to address this gap.^3,6,24^ Before standardization can begin, however, it is first necessary to examine the current literature to understand how authors and practitioners are currently defining stimulability testing and measuring its outcomes. Thus, the current scoping review addressed the following two research questions:

1. How is stimulability testing defined and described in the current voice disorders peer-reviewed literature?
2. How are stimulability testing outcomes defined and described in the current voice disorders peer-reviewed literature?

## Methods

The study followed the five-step methodological framework for scoping reviews,^25,26^ which involves steps to identify the literature (steps 1-3) and analyze and present the results both numerically and thematically (steps 4 and 5).

### Step 1. Identification of the research question

The two research questions guiding this review are as outlined above; namely (1) How is stimulability testing defined and described in the current voice disorders peer-reviewed literature? and (2) How are stimulability testing outcomes defined and described in the current voice disorders peer-reviewed literature?

### Step 2. Identifying the relevant research studies

A literature search was conducted on July 24^th^, 2025 in three search engines; PubMed, EBSCOhost, and Google Scholar. The following search terms were used in PubMed and EBSCOhost: “*(voice OR dysphon*) AND stimulab*”.* The following search string was used for Google Scholar, with the option to search within citing articles turned off: “*voice OR dysphonia OR dysphonic OR vocal OR stimulable “stimulability”-phonological-child”.* No date restrictions were used in the literature search.

While scoping reviews frequently involve textbooks and other grey literature (i.e., non-peer-reviewed literature such as theses and dissertations), the focus of the current study was to examine the highest level of current evidence regarding stimulability testing in voice disorders. In other words, only literature that had been subject to peer review (and thus, in theory, had received critical review of the definitions of stimulability testing procedures) was included.

Additionally, the use of two key words – “stimulability” and “stimulable” – were used to guide the selection of literature and data extraction within literature given the clinical prevalence of this terminology.^3,4^ Given the above, the following inclusion criteria were used to assess the results.

a) Only resources published in the English language available to the author to review were included.
b) Only peer-reviewed articles were included; book chapters, conference proceedings/abstracts, and unpublished theses and dissertations were excluded.
c) Only studies referencing stimulability testing in the context of voice disorders (including hypokinetic dysarthria, Parkinson’s disease or Parkinsonism, or treatments specific to these disorders) were included. Studies referencing stimulability testing in the contexts of speech sound disorders, upper airway disorders, resonance disorders specific to structural changes (i.e., cleft lip and/or palate), hearing disorders, aphasia, or dysarthria that was not specific to Parkinsonism, were excluded.
d) Only articles that used the terms “stimulability” or “stimulable” within the article text were included. Further, these terms had to be used to refer to behavioral measures; articles that referred to physiologic measures of nerve conductance (e.g., “electrophysiologic stimulability”) were excluded.

### Step 3. Study Selection

The literature search process is illustrated in Figure 1. A total of 569 records we e found in the initial literature search. After removal of duplicates from the three databases, 541 unique records remained. Titles were screened for relevance to the inclusion criteria listed in the methods section; 396 articles were removed during this stage, leaving 145 articles for full-text examination. Forty articles were removed for further consideration as they used the terms “stimulability” or “stimulable” to refer to a physiologic measure of nerve conductance. Eleven articles were removed because there was no mention of the terms “stimulability” or “stimulable” within the article text (instead, these terms generally appeared within the references section of excluded articles). Four articles were removed from consideration as they were either not published in English (n = 3) or not accessible for review (n = 1). Two more articles were removed after full-text read throughs, as one mentioned the term “stimulability” only as a direct quote from another article that was previously included, and another was the transcription of a lecture delivered at a conference and was therefore not peer-reviewed despite being published in a peer-reviewed journal. The remaining 88 articles were included in the analysis.

**Figure 1.**
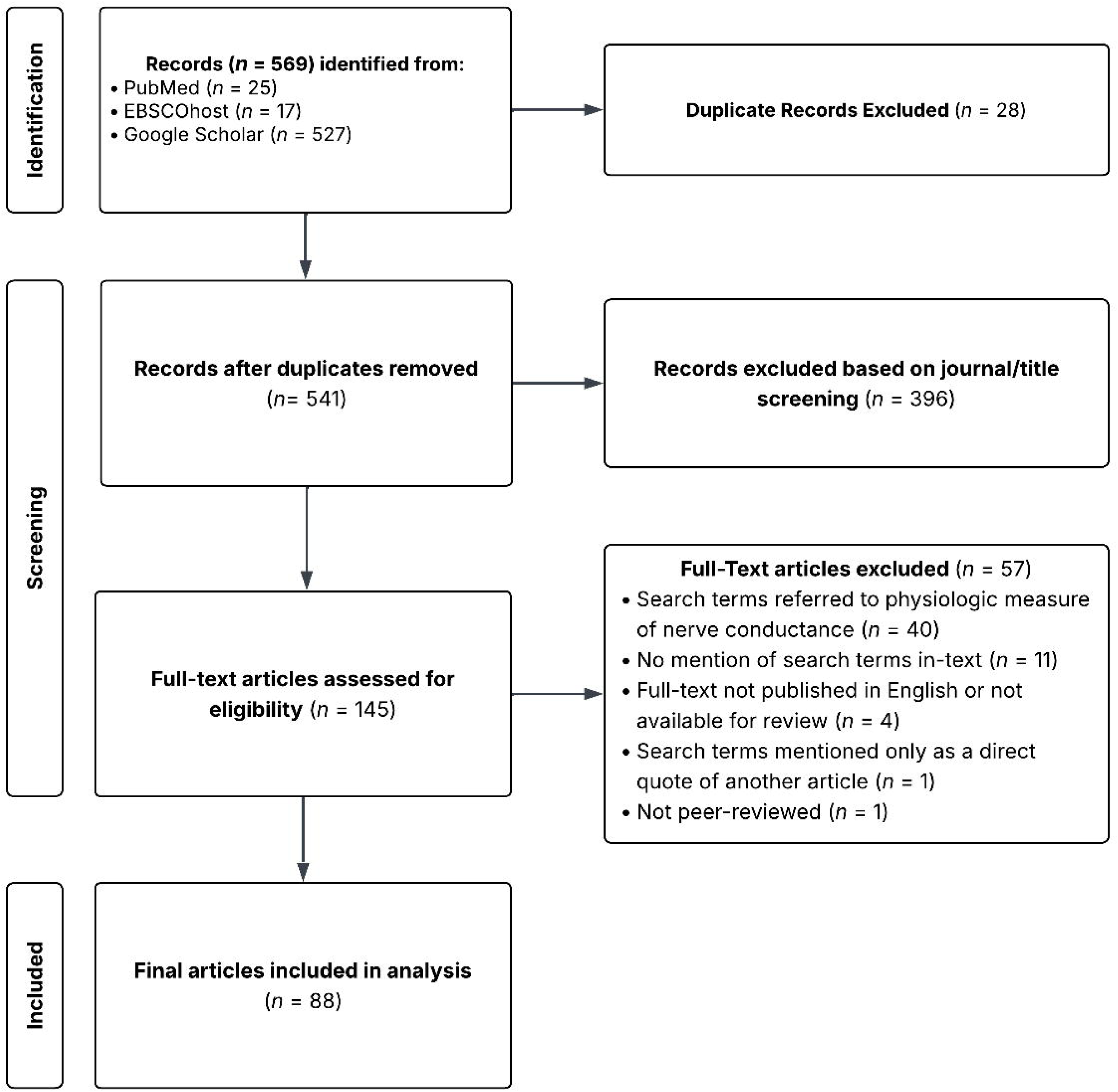
Illustration of search strategy and selection of studies.

### Step 4. Charting the Data

Within each article, the author recorded all instances of the terms “stimulability” and “stimulable” and their surrounding context. Each instance was evaluated to determine if the term referred to stimulability testing or the outcomes of stimulability testing. Definitions of each term, if provided, were recorded. If a definition was not provided, the author extracted relevant surrounding information, such as implied target behaviors (e.g., “stimulable for a better voice”^18(p2)^) or measurement scales used for stimulability testing outcomes (e.g., “highly stimulable”^5(p3)^).

### Step 5. Collating, Summarizing, and Presenting Findings

#### Definitions of Stimulability Testing and Stimulability Testing Outcomes

It became apparent early in the data extraction that, compared to the number of articles retrieved, the number of articles that provided definitions of either stimulability testing or stimulability testing outcomes was relatively small. Because of this, articles were classified based on the level of detail provided when discussing stimulability testing in-text. Articles that provided a definition were classified as “Definition provided” and were further classified on whether they provided a definition for stimulability testing alone, stimulability testing outcomes alone, or both stimulability testing and stimulability testing outcomes. The second classification was “Described rather than defined”; this classification was used when a clinical description stimulability testing or stimulability testing outcomes was provided rather than a higher-level definition of the essential qualities of either aspect. Finally, articles that used search terminology in isolation, without further description or definition of the terminology itself, were classified as “Mentioned only, without a definition”.

Once collected, the author conducted thematic analysis^27^ of the definitions to describe common themes that occurred across definitions. This involved reading of each definition and tagging sections of the definition with a code (i.e., a short phrase capturing the meaning of each section). Codes were then evaluated for broader patterns and clustered into themes based on common conceptual meanings. This process was performed iteratively across each code until the author felt the final the final themes captured all codes, no codes was captured by more than one theme, and each theme was conceptually distinct.

## Results

Publication dates for the included articles are displayed in Figure 2. Article publication dates ranged from 1997 to 2025, with the majority of articles published in 2017 or later. There have been consistent publications with the search terms since 2014, and several years since then have had over 10 publications including the search terms. The year with the most included publications was 2023, which had 16 publications.

**Figure 2.**
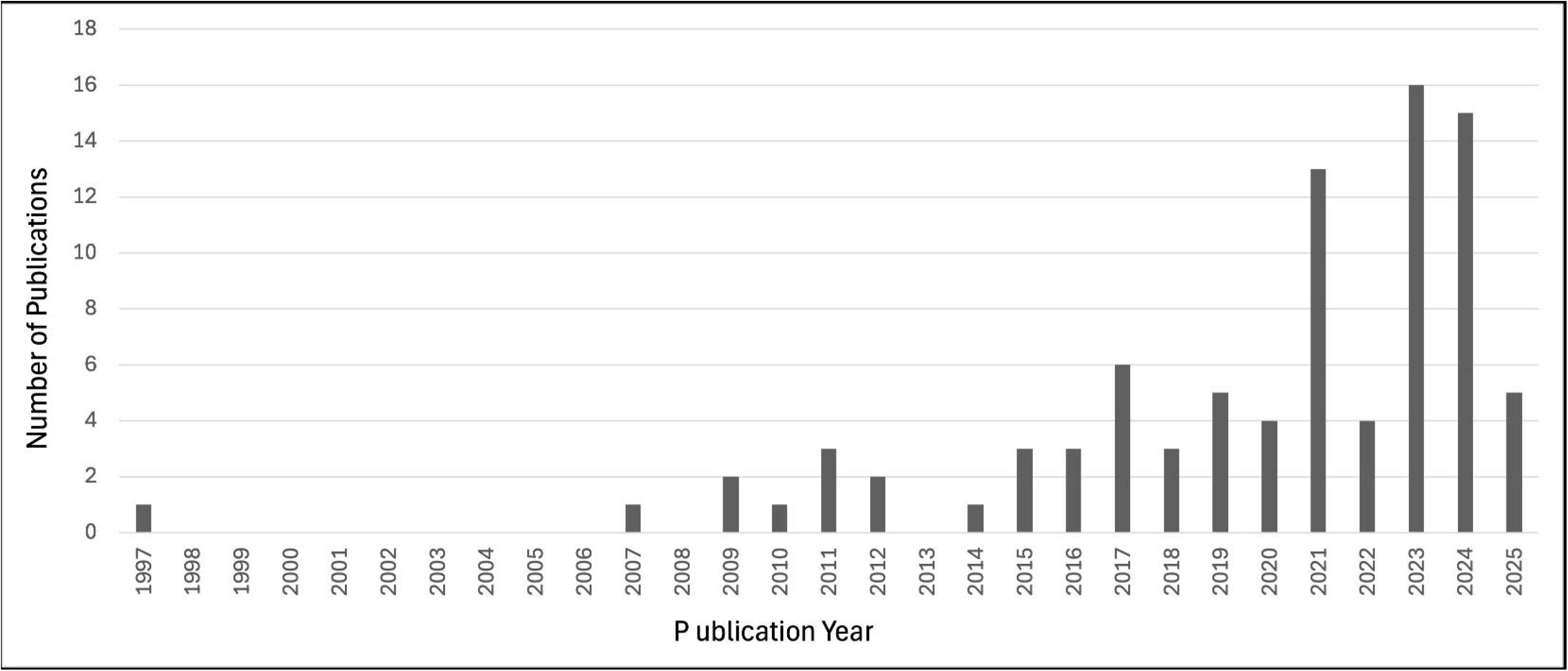
Article publication frequency of included studies.

### Classification of Articles

The classification of definition type (i.e., “Defined”, “Described rather than defined”, and “Mentioned only, without a definition”) for each article are listed in Table 1 and summarized in Table 2. Table 2 also provides a summary of the average number of times the search terms (“stimulability” and “stimulable”) were found within the article text across each classification type.

**Table 1.**
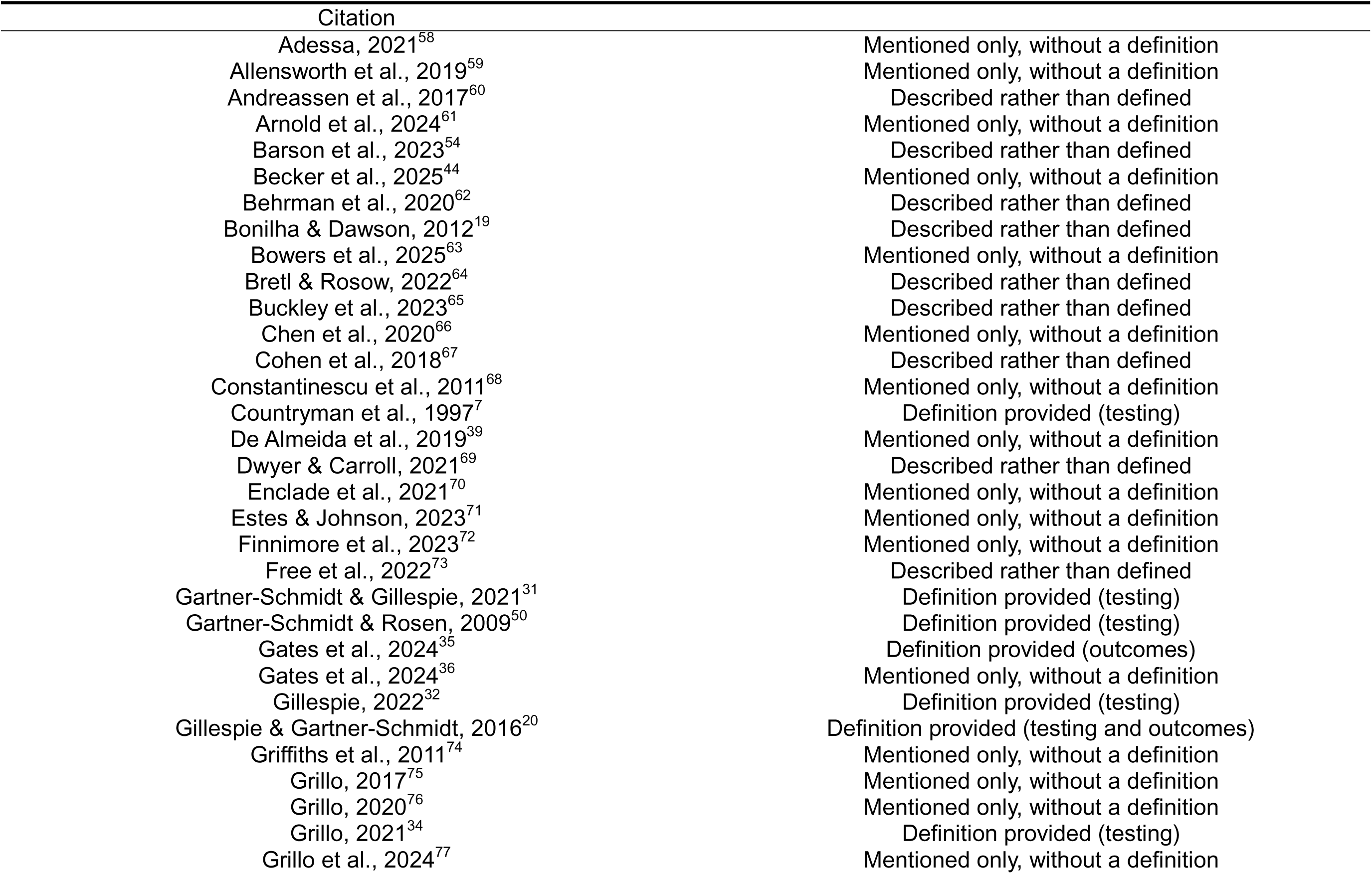
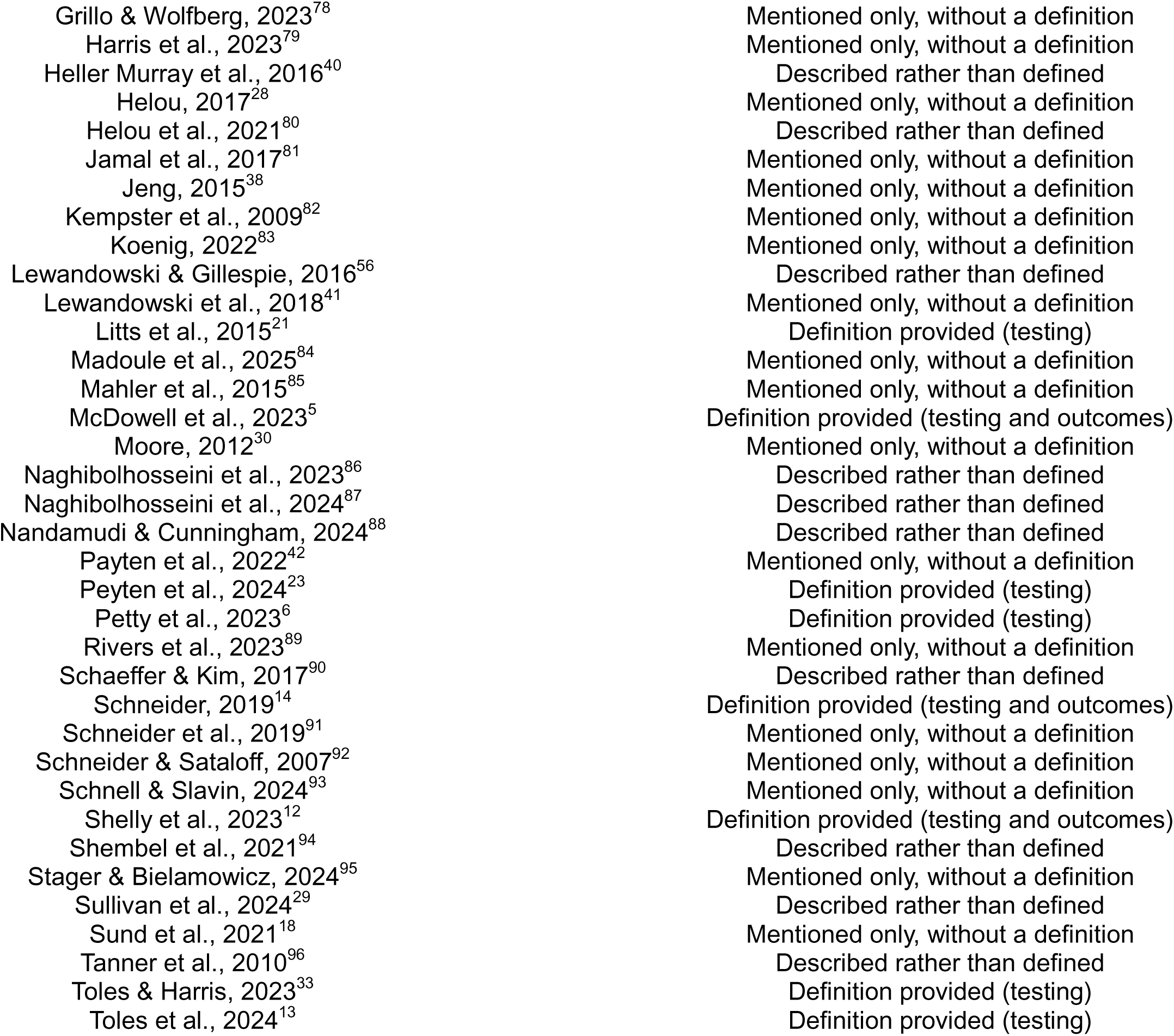
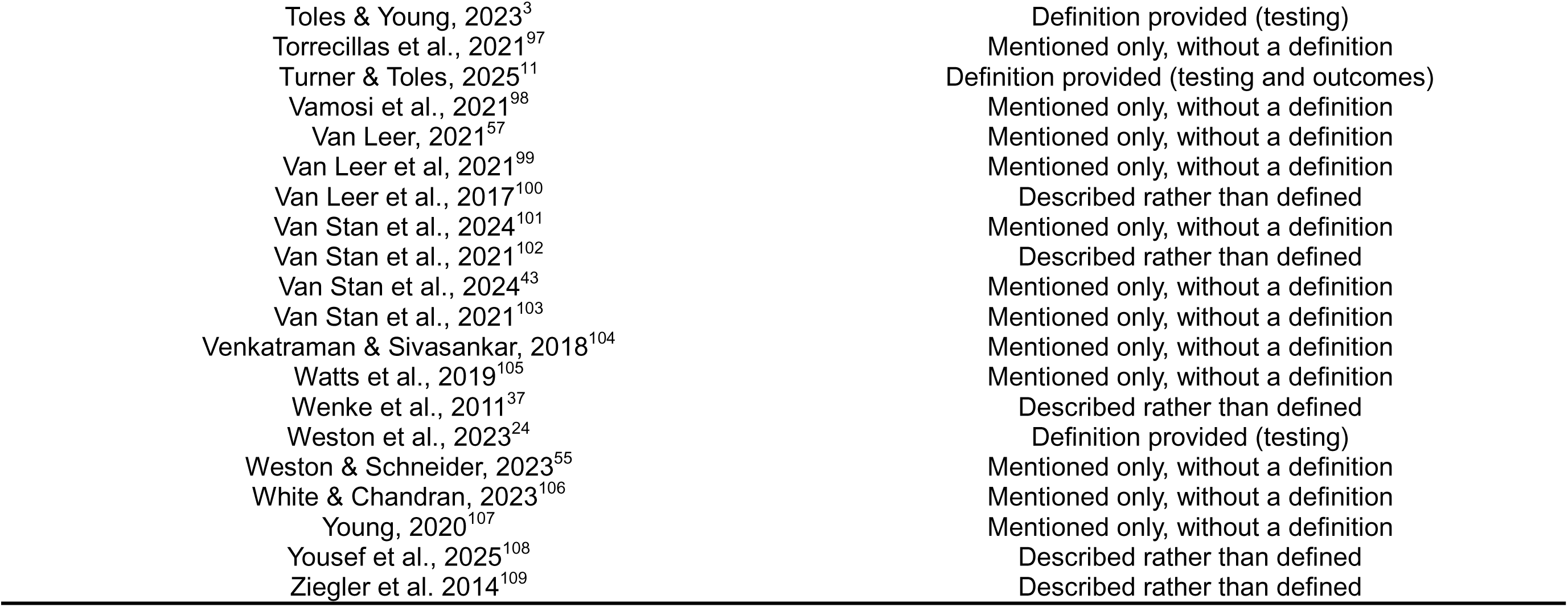
Articles included in the literature review.

| Citation | Classification |
| --- | --- |
| Adessa, 2021<sup>58</sup> | Mentioned only, without a definition |
| Allensworth et al., 2019<sup>59</sup> | Mentioned only, without a definition |
| Andreassen et al., 2017<sup>60</sup> | Described rather than defined |
| Arnold et al., 2024<sup>61</sup> | Mentioned only, without a definition |
| Barson et al., 2023<sup>54</sup> | Described rather than defined |
| Becker et al., 2025<sup>44</sup> | Mentioned only, without a definition |
| Behrman et al., 2020<sup>62</sup> | Described rather than defined |
| Bonilha & Dawson, 2012<sup>19</sup> | Described rather than defined |
| Bowers et al., 2025<sup>63</sup> | Mentioned only, without a definition |
| Bretl & Rosow, 2022<sup>64</sup> | Described rather than defined |
| Buckley et al., 2023<sup>65</sup> | Described rather than defined |
| Chen et al., 2020<sup>66</sup> | Mentioned only, without a definition |
| Cohen et al., 2018<sup>67</sup> | Described rather than defined |
| Constantinescu et al., 2011<sup>68</sup> | Mentioned only, without a definition |
| Countryman et al., 1997<sup>7</sup> | Definition provided (testing) |
| De Almeida et al., 2019<sup>39</sup> | Mentioned only, without a definition |
| Dwyer & Carroll, 2021<sup>69</sup> | Described rather than defined |
| Enclade et al., 2021<sup>70</sup> | Mentioned only, without a definition |
| Estes & Johnson, 2023<sup>71</sup> | Mentioned only, without a definition |
| Finnimore et al., 2023<sup>72</sup> | Mentioned only, without a definition |
| Free et al., 2022<sup>73</sup> | Described rather than defined |
| Gartner-Schmidt & Gillespie, 2021<sup>31</sup> | Definition provided (testing) |
| Gartner-Schmidt & Rosen, 2009<sup>50</sup> | Definition provided (testing) |
| Gates et al., 2024<sup>35</sup> | Definition provided (outcomes) |
| Gates et al., 2024<sup>36</sup> | Mentioned only, without a definition |
| Gillespie, 2022<sup>32</sup> | Definition provided (testing) |
| Gillespie & Gartner-Schmidt, 2016<sup>20</sup> | Definition provided (testing and outcomes) |
| Griffiths et al., 2011<sup>74</sup> | Mentioned only, without a definition |
| Grillo, 2017<sup>75</sup> | Mentioned only, without a definition |
| Grillo, 2020<sup>76</sup> | Mentioned only, without a definition |
| Grillo, 2021<sup>34</sup> | Definition provided (testing) |
| Grillo et al., 2024<sup>77</sup> | Mentioned only, without a definition |
| Grillo & Wolfberg, 2023<sup>78</sup> | Mentioned only, without a definition |
| Harris et al., 2023<sup>79</sup> | Mentioned only, without a definition |
| Heller Murray et al., 2016<sup>40</sup> | Described rather than defined |
| Helou, 2017<sup>28</sup> | Mentioned only, without a definition |
| Helou et al., 2021<sup>80</sup> | Described rather than defined |
| Jamal et al., 2017<sup>81</sup> | Mentioned only, without a definition |
| Jeng, 2015<sup>38</sup> | Mentioned only, without a definition |
| Kempster et al., 2009<sup>82</sup> | Mentioned only, without a definition |
| Koenig, 2022<sup>83</sup> | Mentioned only, without a definition |
| Lewandowski & Gillespie, 2016<sup>56</sup> | Described rather than defined |
| Lewandowski et al., 2018<sup>41</sup> | Mentioned only, without a definition |
| Litts et al., 2015<sup>21</sup> | Definition provided (testing) |
| Madoule et al., 2025<sup>84</sup> | Mentioned only, without a definition |
| Mahler et al., 2015<sup>85</sup> | Mentioned only, without a definition |
| McDowell et al., 2023<sup>5</sup> | Definition provided (testing and outcomes) |
| Moore, 2012<sup>30</sup> | Mentioned only, without a definition |
| Naghibolhosseini et al., 2023<sup>86</sup> | Described rather than defined |
| Naghibolhosseini et al., 2024<sup>87</sup> | Described rather than defined |
| Nandamudi & Cunningham, 2024<sup>88</sup> | Described rather than defined |
| Payten et al., 2022<sup>42</sup> | Mentioned only, without a definition |
| Peyten et al., 2024<sup>23</sup> | Definition provided (testing) |
| Petty et al., 2023<sup>6</sup> | Definition provided (testing) |
| Rivers et al., 2023<sup>89</sup> | Mentioned only, without a definition |
| Schaeffer & Kim, 2017<sup>90</sup> | Described rather than defined |
| Schneider, 2019<sup>14</sup> | Definition provided (testing and outcomes) |
| Schneider et al., 2019<sup>91</sup> | Mentioned only, without a definition |
| Schneider & Sataloff, 2007<sup>92</sup> | Mentioned only, without a definition |
| Schnell & Slavin, 2024<sup>93</sup> | Mentioned only, without a definition |
| Shelly et al., 2023<sup>12</sup> | Definition provided (testing and outcomes) |
| Shembel et al., 2021<sup>94</sup> | Described rather than defined |
| Stager & Bielamowicz, 2024<sup>95</sup> | Mentioned only, without a definition |
| Sullivan et al., 2024<sup>29</sup> | Described rather than defined |
| Sund et al., 2021<sup>18</sup> | Mentioned only, without a definition |
| Tanner et al., 2010<sup>96</sup> | Described rather than defined |
| Toles & Harris, 2023<sup>33</sup> | Definition provided (testing) |
| Toles et al., 2024<sup>13</sup> | Definition provided (testing) |
| Toles & Young, 2023<sup>3</sup> | Definition provided (testing) |
| Torrecillas et al., 2021<sup>97</sup> | Mentioned only, without a definition |
| Turner & Toles, 2025<sup>11</sup> | Definition provided (testing and outcomes) |
| Vamosi et al., 2021<sup>98</sup> | Mentioned only, without a definition |
| Van Leer, 2021<sup>57</sup> | Mentioned only, without a definition |
| Van Leer et al, 2021<sup>99</sup> | Mentioned only, without a definition |
| Van Leer et al., 2017<sup>100</sup> | Described rather than defined |
| Van Stan et al., 2024<sup>101</sup> | Mentioned only, without a definition |
| Van Stan et al., 2021<sup>102</sup> | Described rather than defined |
| Van Stan et al., 2024<sup>43</sup> | Mentioned only, without a definition |
| Van Stan et al., 2021<sup>103</sup> | Mentioned only, without a definition |
| Venkatraman & Sivasankar, 2018<sup>104</sup> | Mentioned only, without a definition |
| Watts et al., 2019<sup>105</sup> | Mentioned only, without a definition |
| Wenke et al., 2011<sup>37</sup> | Described rather than defined |
| Weston et al., 2023<sup>24</sup> | Definition provided (testing) |
| Weston & Schneider, 2023<sup>55</sup> | Mentioned only, without a definition |
| White & Chandran, 2023<sup>106</sup> | Mentioned only, without a definition |
| Young, 2020<sup>107</sup> | Mentioned only, without a definition |
| Yousef et al., 2025<sup>108</sup> | Described rather than defined |
| Ziegler et al. 2014<sup>109</sup> | Described rather than defined |

**Table 2.** Summary of Article Classifications.

| Article Classification | Count (percent) | Mean (SD) of search term mentions in article text<sup>1</sup> |
| --- | --- | --- |
| Search Terms Mentioned Only, Without a Definition | 46 (52%) | 2.10 (3.66) |
| Search Terms Described Rather than Defined | 24 (27%) | 4.50 (9.33) |
| Definition Provided for Stimulability Testing Only | 12 (14%) | 29.30 (54.22) |
| Definition Provided for Stimulability Testing Outcomes Only | 1 (1%) | 23.00 (n/a) |
| Definition Provided for Stimulability Testing and Stimulability Testing Outcomes | 5 (6%) | 72.20 (57.46) |
| Total | 88 (100%) | 10.69 (29.50) |

<sup>1</sup>Search terms included “stimulability” and “stimulable”.

The most common classification type was “Mentioned only, without a definition”, which described over half (52%) of included articles. As seen in Table 2, these articles infrequently mentioned search terms, and generally did so isolation without further description. For example, one article used a search term only once in the following context: “Further, a good voice therapist pulls tricks out of their sleeve, seeking moments of stimulability until something works and sticks.”^28(p86)^ Over a quarter (27%) of articles provided a clinical description of what was occurring during or after stimulability testing rather than a higher-level definition of the essential qualities of stimulability testing or stimulability testing outcomes (i.e., Described rather than defined). For example, Sullivan et al.^29^ described stimulability testing by stating it was “conducted during the initial evaluation by instructing the participants to repeat speech tasks using intent following a clinician model.”^(p1934)^ While this description provides insight into how stimulability testing may be performed, it fails to provide detail regarding what stimulability testing is and how it differs from other aspects of the behavioral voice evaluation. Twenty percent of articles provided a definition of stimulability testing or stimulability testing outcomes, although most frequently articles provided a definition of stimulability testing alone. Articles classified as providing definitions used search terms significantly more often than articles in other categories, as measured by a Welch’s t-test (*t*(17) = 2.90, *p* = 0.01).

Regardless of article classification type, the use of search terms within each article was analyzed to determine whether the authors were discussing stimulability testing or stimulability testing outcomes. Of the included articles, 33 (38%) discussed stimulability testing only, while 24 (27%) discussed stimulability testing outcomes only. Thirty articles (34%) discussed both stimulability testing and stimulability testing outcomes. There was one instance where the use of the search terms was ambiguous to the point where it was indeterminate whether the term “stimulability” referred to stimulability testing or stimulability testing outcomes: “For other clients the process of individualized prescription of yoga (including ‘*stimulability’* [italics added], the use of self-generated internal imagery, and on-going monitoring of responses) ensured that yoga practices were modified until a state of ‘flow’ could be achieved.“ ^30(pp146-147)^

### Definitions of Stimulability Testing

Seventeen of the 88 included articles (19%) provided definitions of stimulability testing, which are displayed in Table 3. Thematic analysis of the definitions resulted in five themes: (1) Modifying Behaviors, (2) Assessing Patient Response, (3) Using Facilitators, (4) Specifying Goals, and (5) Contextualizing the Evaluation. Representative quotes from each theme are displayed in Table 4.

**Table 3.** Definitions of Stimulability Testing Provided by Articles within Literature Search.

| Citation | Definition of Stimulability Testing |
| --- | --- |
| Countryman et al. (1997)<sup>7</sup> | "Stimulability testing is recommended during pretreatment screening to determine if the individual is capable of producing a louder voice easily and without straining." (p. 82) |
| Gartner-Schmidt & Gillespie (2021)<sup>31</sup> | "Stimulability assessment refers to the evaluation of a patient's ability to modify a behavior when provided models." (p. 38) |
| Gartner-Schmidt & Rosen (2009)<sup>50</sup> | "Patients are evaluated for their ability to either feel and/or sound better using appropriate voice balancing techniques (i.e., vocal stimulability/vocal plasticity)." (p. 8) |
| Gillespie (2022)<sup>32</sup> | "P.L.'s voice was assessed for ability to change in response to therapeutic tasks" (p. 714) |
| Gillespie & Gartner-Schmidt (2016)<sup>20</sup> | " Stimulability assessment refers to the evaluation of an individual's ability to modify a behavior when provided with models or cues" (p. 507.e15) |
| Grillo (2021)<sup>34</sup> 5/21/2026 2:43:00 PM | "Stimulability testing involved finding the best voice production techniques that facilitate the "new" voices." (p 569) |
| Litts et al. (2015)<sup>21</sup> | " Stimulability testing refers to the evaluation of an individual's ability to modify a behavior when provided with models or cues." (p. 2140) |
| McDowell et al. (2023)<sup>5</sup> | "Voice stimulability is defined as the assessment of a person's ability to make immediate changes to their voice." (p. 1) |
| Payten et al. (2024)<sup>23</sup> | "...observation of the person's ability to modify the voice to direction (voice therapy stimulability testing)." (p 2794) |
| Petty et al. (2023)<sup>6</sup> | " Stimulability is the assessment of an individual's ability to modify a behavior when provided with models or cues." (p. 3) |
| Schneider (2019)<sup>14</sup> | "During vocal stimulability testing, therapeutic tools are used by the clinician to determine the patient's ability to follow vocal direction and produce a change in ease and/or quality of voice. The patient's self-perception of changes in the sound and feel is also assessed." (p. 475) |
| Shelly et al. (2023)<sup>12</sup> | " This trial therapy is also known as stimulability testing. Stimulability refers to the ability to immediately change a behavior when provided with a model, prompt, or cue." (p. 1) |
| Toles & Harris (2023)<sup>33</sup> | "In the voice evaluation, the SLP will perform voice stimulability testing, in which the clinician assesses the patient's ability to modify the voice when provided with different facilitating techniques and cues." (p. 56-57). "Stimulability testing is a dynamic process that can serve to identify how easily, and with what certain techniques, a patient can manipulate the voice to achieve a more balanced production." (p. 322) |
| Toles & Young (2023)<sup>3</sup> | " Voice stimulability is defined as the ability to modify the voice when provided with demonstration and/or cues." (p. 1) |
| Toles et al. (2024)<sup>13</sup> | "The primary goal of stimulability testing is to determine whether a patient can immediately change their vocal quality, effort, or efficiency given a variety of facilitating tasks and levels of cueing" (p. 1) |
| Turner & Toles (2025)<sup>11</sup> | "Stimulability testing, or the assessment of an individual's ability to change their behavior in the presence of cues and supports, is widely used by voice-specialized speech-language pathologists during initial voice evaluation to guide treatment decisions and estimate prognosis for improvement." (p 1) |
| Weston et al. (2023)<sup>24</sup> | " Stimulability testing is a critical component of a voice evaluation to assess the patient's ability to make vocal changes related to their voice goal, determine readiness for change, and establish self-efficacy through a mastery experience." (p. 1) |

**Table 4.** Results from Thematic Analysis of Stimulability Testing Definitions.

| Theme | Supporting Quotes |
| --- | --- |
| Modifying Behaviors | "...modify a behavior..."<sup>31</sup><br>"...make vocal changes..."<sup>24</sup><br>"...feel and/or sound better..."<sup>50</sup><br>"...self-perception of changes in the sound and feel..."<sup>14</sup> |
| Assessing Patient Response | "Patients are evaluated for their ability to..."<sup>50</sup><br>"a dynamic process that can serve to identify how easily... a patient can manipulate..."<sup>33</sup><br>"...determine if the individual is capable of producing..."<sup>7</sup><br>"...an individual's ability to modify..."<sup>21</sup> |
| Using Facilitators | "...using appropriate voice balancing techniques..."<sup>50</sup><br>"...when provided with a model, prompt, or cue..."<sup>12</sup><br>"...when provided with different facilitating techniques and cues..."<sup>33</sup><br>"...given a variety of facilitating tasks and levels of cueing..."<sup>13</sup> |
| Specifying Goals | "...finding the best voice production techniques..."<sup>34</sup><br>"Stimulability testing... is widely used... to guide treatment decisions and estimate prognosis for improvement..."<sup>11</sup><br>"...determine readiness for change, and establish self-efficacy through a mastery experience..."<sup>24</sup> |
| Contextualizing the Evaluation | "Stimulability testing is recommended during pretreatment screening..."<sup>7</sup><br>"...make immediate changes to their voice..."<sup>5</sup><br>"Stimulability testing is a dynamic process..."<sup>33</sup><br>"...a critical component of a voice evaluation..."<sup>24</sup> |

In the first theme, Modifying Behaviors, definitions described eliciting a behavioral change from the participant. A wide variety of behaviors were the target of modification; some definitions did not specify a specific behavior, ^6,11,12,20,21,31^ others specified a broad behavioral category such as “voice”^3,5,13,23,32^; and still others specified a very specific vocal behavior (e.g., “louder voice without strain”^7(p82)^). Despite this, the Modifying Behaviors theme was apparent in all seventeen definitions examined within this review, suggesting a strong conceptual cohesion between this theme and stimulability testing. In the second theme, Assessing Patient Response, definitions described evaluating the patients’ response to elicited changes. Wording used by authors suggested patients would vary in their response to stimulability testing (e.g., “a patient’s *ability* [italics added] to modify a behavior”^31(p38)^, “identify *how easily* [italics added]… a patient can manipulate the voice…”^33(p322)^). Fifteen of the seventeen (88%) of definitions included codes that were categorized under this theme. Within the third theme, Using Facilitators, definitions specified that the use of facilitating techniques, such as modeling, cueing, or other therapeutic supports as an aspect of stimulability testing. Most definitions (14/17; 82%) described the use of some form of facilitator, most commonly “cues” (8/14; 57%) or “models” (5/14; 36%).

Within the Specifying Goals theme, definitions discussed the various goals of performing stimulability testing, such as guiding treatment decisions^11,34^ and establishing patient self-efficacy^24^. Four of the seventeen (24%) definitions included codes within this theme. Finally, within the Contextualizing Evaluation theme, definitions included information such as when stimulability testing occurred (i.e., during pretreatment screening^7^ or the voice evaluation^11^) and the expected time course of behavioral changes during stimulability testing (e.g., immediate change^5,12,13^). Six definitions (35%) included codes that were classified within the Contextualizing Evaluation theme.

### Definitions of Stimulability Testing Outcomes

Seven articles (8%) provided definitions of stimulability testing outcomes; two of these articles provided multiple definitions, yelling a total of ten definitions for analysis. These definitions are displayed in Table 5. Three themes emerged from the thematic analysis of these definitions: (1) Defining Outcome (2) Quantifying Change, and (3) Supporting Change. The Quantifying Change included two subthemes, *Change Measurement* and *Change Context*.

**Table 5.**
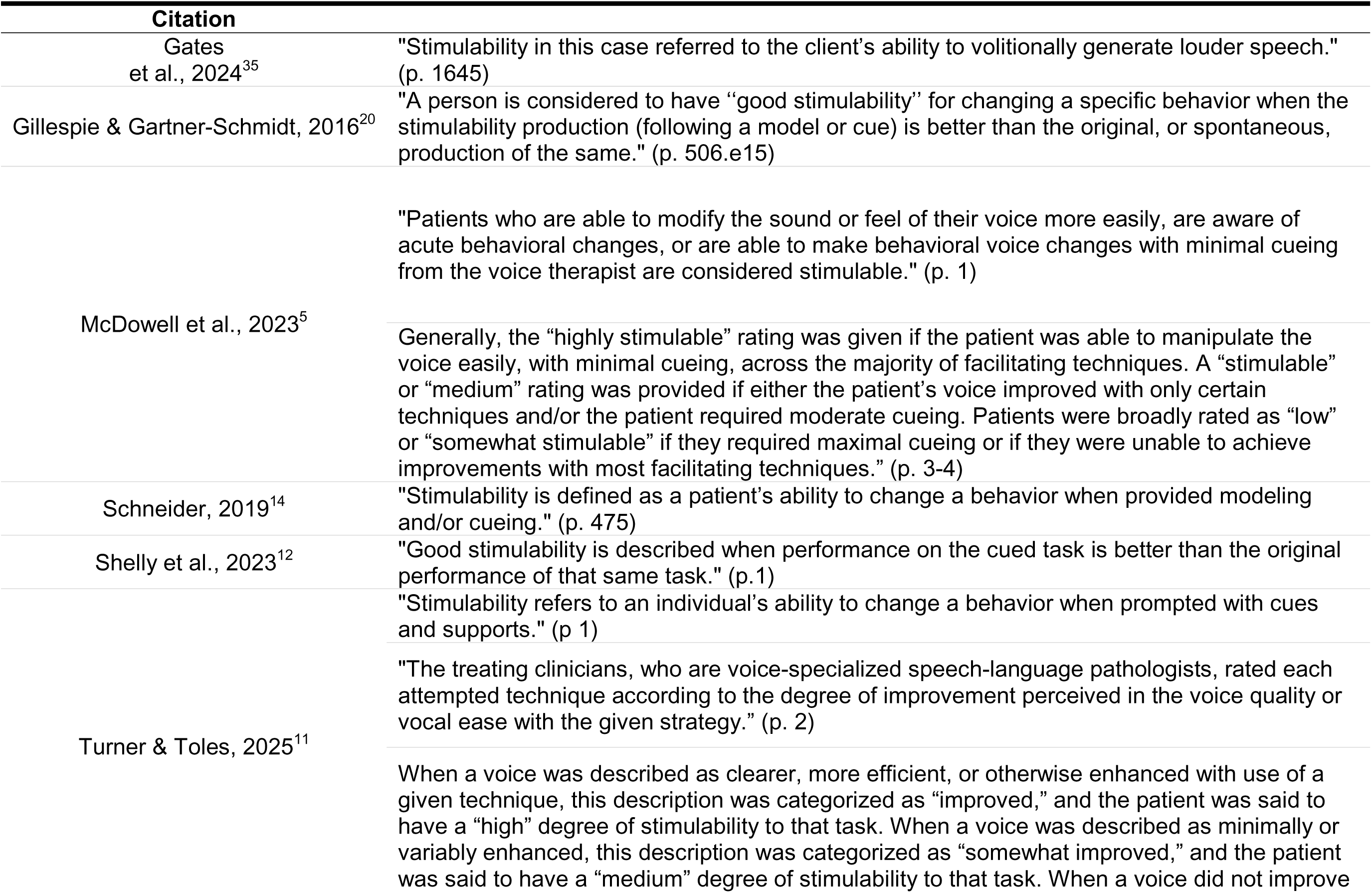
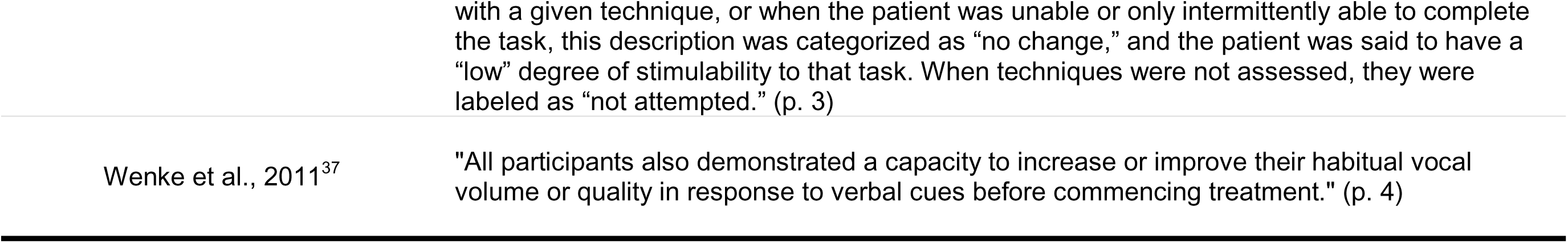
Definitions and Analysis for Stimulability Testing Outcomes.

These themes and subthemes, along with supporting quotes, are displayed in Table 6.

**Table 6.** Results of Thematic Analysis for Stimulability Testing Outcomes.

| Theme | Subthemes | Supporting Quotes |
| --- | --- | --- |
| Defining Outcomes | n/a | "...generate louder speech."<sup>35</sup><br>"...patients who... are aware of acute behavioral changes..."<sup>5</sup><br>"...the degree of improvement perceived in the voice quality or vocal ease..."<sup>11</sup><br>"...performance on the cued task..."<sup>12</sup><br>"...changing a specific behavior..."<sup>20</sup> |
| Quantifying Change | Change Measurement | " a person was considered to have 'good stimulability' ..." <sup>12</sup><br>"the 'highly stimuable' rating was given..." <sup>5</sup><br>"Stimulability in this case referred to..." <sup>35</sup> |
|  | Change Context | "... if the patient was able to manipulate the voice... across the majority of facilitating techniques..." <sup>5</sup><br>"...performance on the cued task is better than the original performance on that same task." <sup>12</sup><br>"the degree of improvement... with the given strategy." <sup>11</sup> |
| Supporting Change | n/a | "...volitionally generate..." <sup>35</sup><br>"...with minimal cueing..." <sup>5</sup><br>"...when prompted with cues and supports." <sup>11</sup><br>"...in response to verbal cues..." <sup>37</sup> |

Within the Defining Outcome theme, descriptions were provided of the outcome (or outcomes) of interest to the clinician during stimulability testing. The number of outcomes described within a definition varied; while all definitions provided at least one outcome, other definitions provided up to six.^11^ The Quantification of Change theme described how meaningful differences in the outcomes were measured, both in terms of the necessary context for measuring change (*Change Context* subtheme) and rating scale types for measuring change (*Change Measurement* subtheme). In the *Change Context* subtheme, definitions provided relational language to describe the behaviors against which change was measured. The most common context specified was comparing a patient’s baseline performance to their performance with a facilitating technique,^11,12,20^ although relative performance on multiple facilitators^5^ was also noted. Six of the 10 (60%) definitions across four articles^5,11,12,35^ had codes within the *Change Context* subtheme. The *Change Measurement* subtheme described the measurement scales used to quantify changes in the outcome measure(s). Eight (80%) of definitions across six articles^5,11,12,14,20,36^ had codes within this subtheme; three (30%) used a binary rating scale (e.g., “good stimulability”, “stimulable”),^5,12,20^ two (20%) used a categorical rating scale (e.g., “highly stimulable”, “somewhat stimulable”),^5,11^ and three (30%) did not specify the rating scale type (e.g., “stimulability”).^11,14,35^

Finally, within the Supporting Change theme, the degree and type of direct clinician support utilized to achieve stimulability testing outcomes were outlined. All definitions across all articles included language that was coded under the Supporting Change theme. The majority of definitions used relatively vague descriptions of support such as “a model or cue”^5,12,14,20^; however, two articles specified the amount of cueing (“minimal cueing”^5^) and one specified the type of cueing (“verbal cues”^37^).

### Other Extracted Information

While many articles did not formally define stimulability testing or stimulability testing outcomes, information surrounding the search terms often resulted in extractable information that could be further examined. The target behavior of stimulability testing was frequently extractable from both descriptions of stimulability testing (e.g., “the SLP had conducted stimulability probes for increased vocal loudness”^38^) and stimulability testing outcomes (e.g., “stimulability to flow phonation”^31^). The type of measure scales used to measure stimulability testing outcomes (e.g., “highly stimulable”^11^) was also frequently extractable. Each of these will be examined in turn below.

### Behavioral Targets of Stimulability Testing

Table 7 outlines the behavioral targets of stimulability testing extracted across 54 articles (61%). Behavioral targets in Table 7 were classified as behavior-specific (either facilitators, such as “resonant voice”^39^, or general behaviors, such as “improved voice quality”^40^) or individual-specific (e.g., “behavioral treatment”^41^). Of the articles with extractable behavioral targets, forty (74%) described behavior-specific targets, with 15 (28%) describing facilitators (e.g., “humming”, “negative practice”, “using intent”), and 25 (46%) describing a general behavior (e.g., “voice function”, “voice change” “voicing efficiency”). Three articles (6%) described individual-specific targets (“behavioral change”, “voice therapy”). Twenty percent of articles (*n* = 11) described targets from multiple categories, most frequently both behavior and individual-specific (e.g., “therapy” and “loud voice”^36^).

**Table 7.**
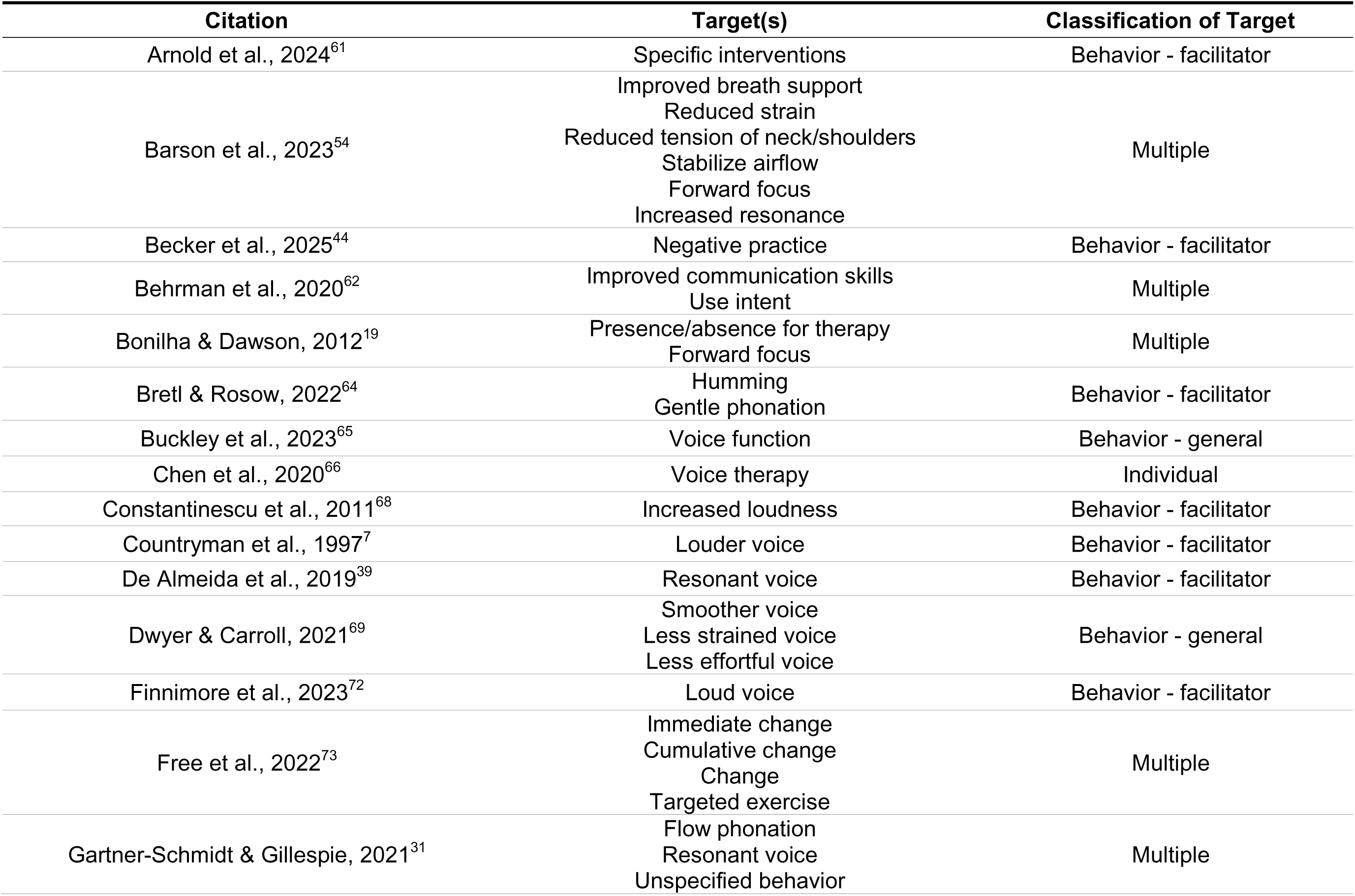
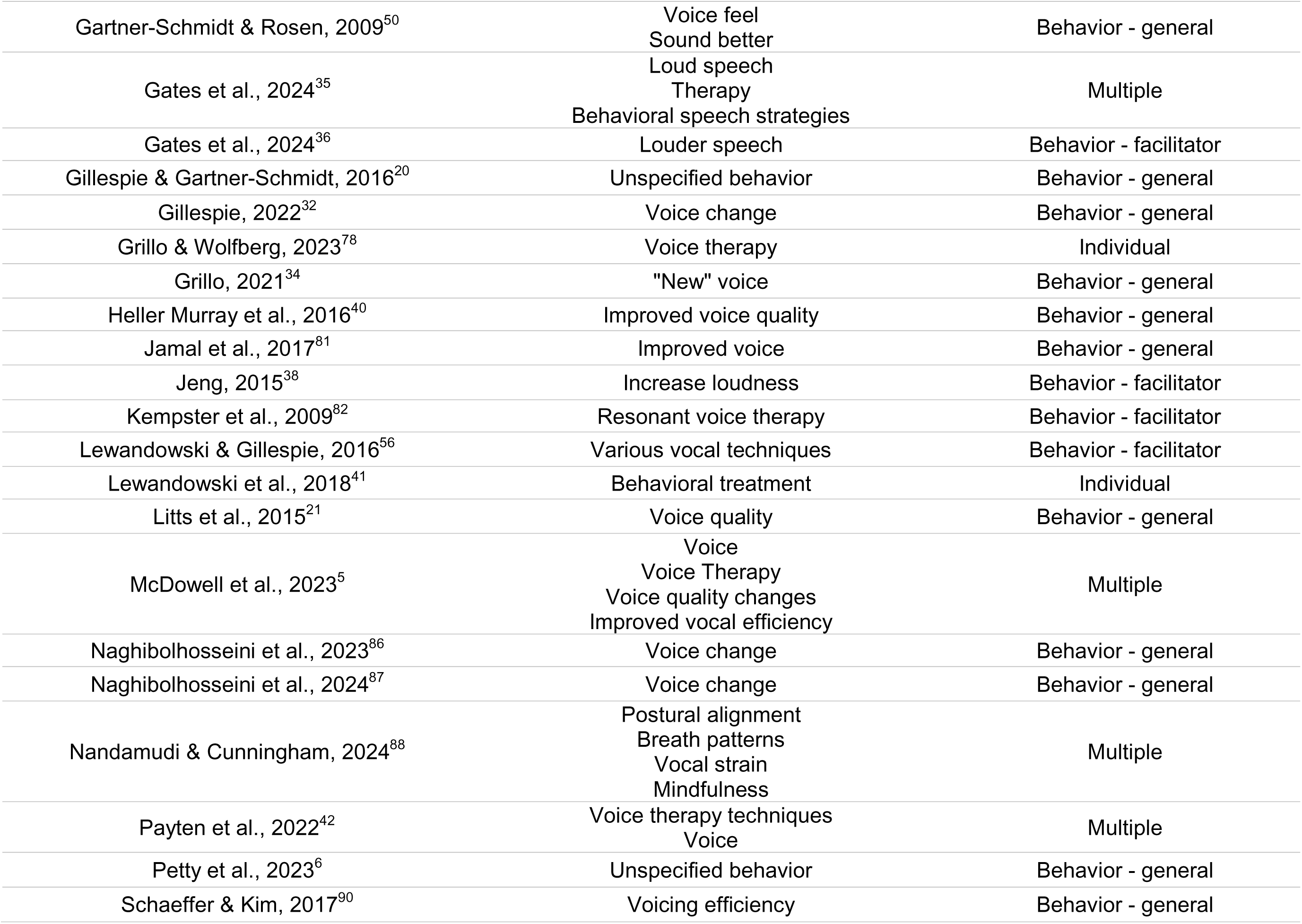
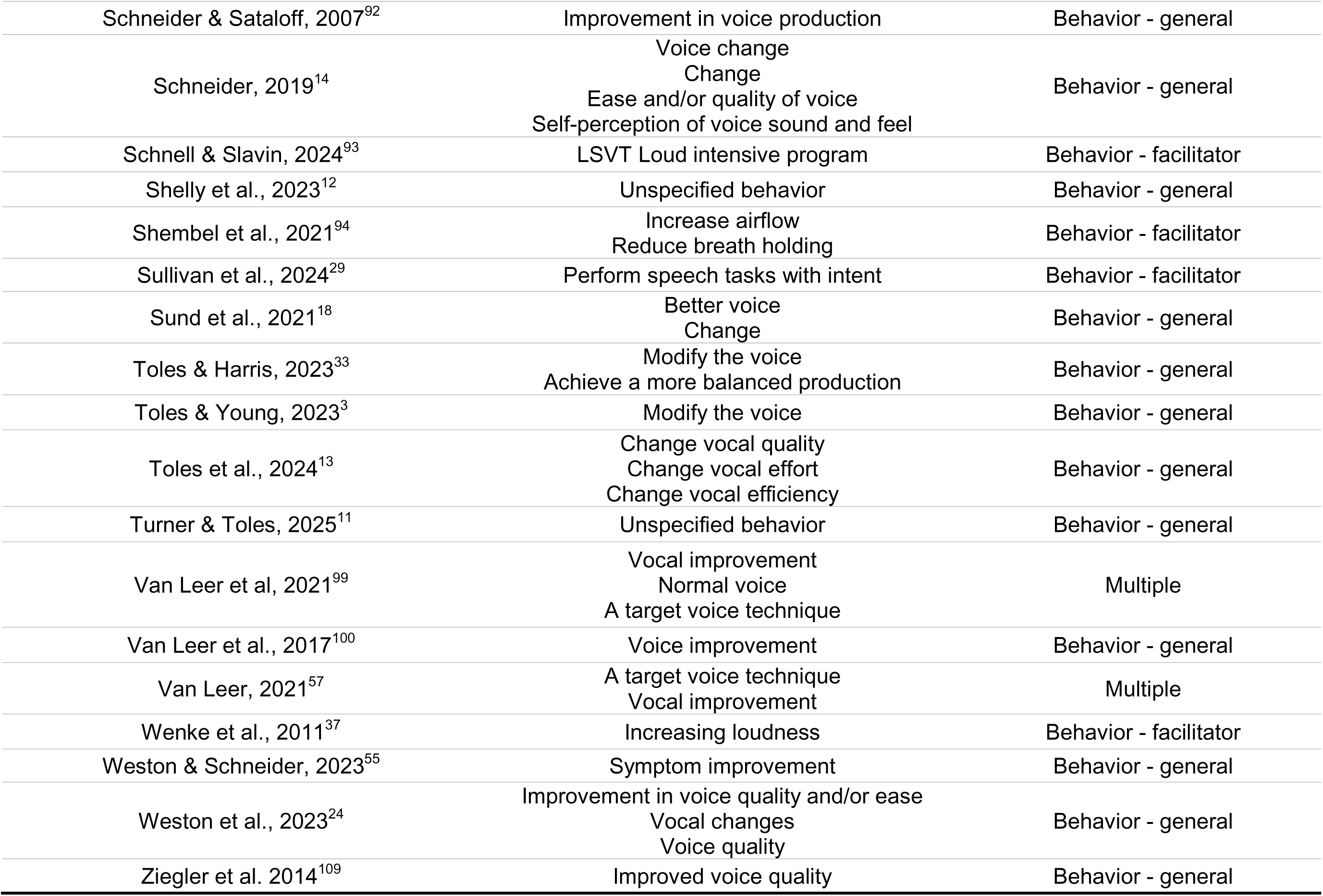
Behavioral Target(s) of Stimulability Testing.

### Measurement Scale Types for Stimulability Testing Outcomes

The types of measurement scales used for stimulability testing outcomes were extractable from the text in 26 of the 54 articles (48%) describing stimulability testing outcomes. These results are displayed in Table 8. Based on the terminology used in each of these articles, the type of measurement scale utilized was classified as either binary (e.g., “stimulable vs not stimulable”^13^), categorical (e.g., “Excellent, good, poor”^42^), or “multiple” if more than one measurement scale type was used. Twenty-one of these articles (81%) used a measurement scale that was classified as binary, and three articles (12%) used a categorical measurement scale.^11,42,43^ Two articles (8%) used multiple measurement scales.^5,44^

**Table 8.**
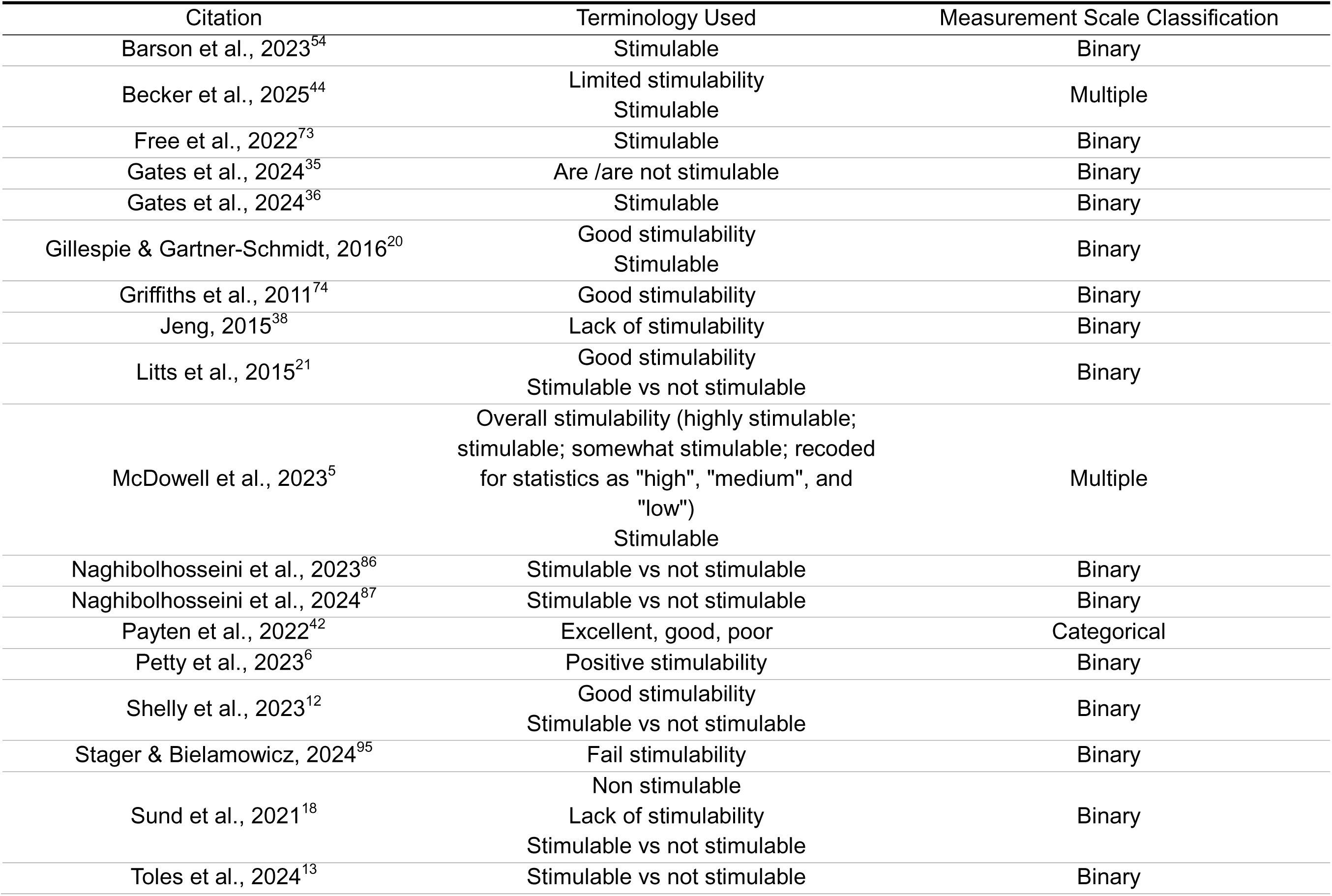
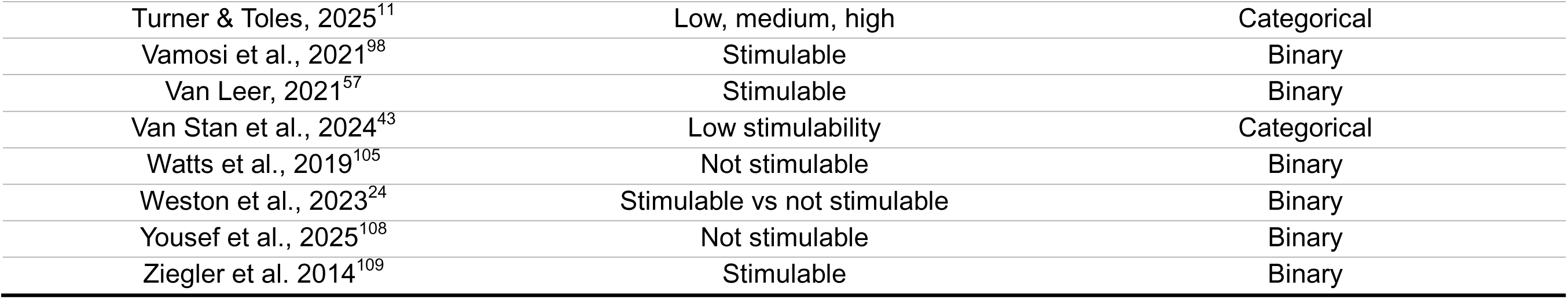
Stimulability Testing Outcome Measurement Scales.

## Discussion

Stimulability testing has a history as an assessment tool for speech-language pathologists specializing in many different areas, including voice disorders. However, current clinical stimulability testing practices vary widely, with less than 15% of practicing speech-language pathologists and laryngologists reporting a standardized stimulability testing procedure in their current practice location.^3,4^ Because of this, definitions of stimulability testing vary widely in the established literature. Thus, the current scoping review examined the definitions provided for stimulability testing as well as the outcomes of stimulability testing in peer-reviewed literature focusing on voice disorders.

### Frequency of Publication

Despite not implementing a date restriction during the literature search, the earliest publication in the current review was from 1997. This was somewhat surprising given the term “stimulability” has been known to be in use within the speech-language pathology literature since the 1950s^1,45,46^ and within voice disorder textbooks since the 1970s.^2,47–49^ The results of the current review suggest that the migration of the terms “stimulable” and “stimulability” to peer-reviewed literature for voice disorders did not begin until the late 20^th^ and early 21^st^ century.

Before this point, these terms may have been more limited in use or used primarily within clinical context. Alternately, different terms may have been used for similar practices; this possibility is discussed further in the limitations section. Publication frequency regarding stimulability testing has increased exponentially within the last decade, likely indicating an increased collective awareness of stimulability testing as a clinical and research tool. As publication frequency increases, however, there is an even greater need to thoughtfully and thoroughly consider how stimulability testing and stimulability outcomes are defined and described within the literature.

### Classification of Articles

The majority of articles included in this review (80%) did not formally define either stimulability testing or stimulability testing outcomes. Instead, more than half of included articles (52%) used the search terms “stimulability” or “stimulable” without any definition or description. Together, these findings suggest a tendency to assume a shared understanding of the meaning of stimulability testing and stimulability testing outcomes amongst readerships. However, as will be further discussed in the following sections, there is no common, consensus definition of stimulability testing nor stimulability testing outcomes in the voice disorders literature. Thus, this review highlights a problematic disconnect between the usage of terminology surrounding stimulability testing compared to the availability of consensus of definitions for such terms. This disconnect undermines the ability of both researchers and clinicians to communicate effectively regarding stimulability testing and stimulability testing outcomes.

### Definitions of Stimulability Testing

The study’s first research question was to examine how stimulability testing is defined and described within the current peer-reviewed literature. Of the 88 articles reviewed, only 17 (19%) provided a definition of stimulability testing. Additionally, more than 70% of articles with a definition of stimulability testing were published within the last five years. This suggests emerging interest within the literature in defining terminology surrounding stimulability testing practices, despite evidence suggesting longer-term use of these terms. However, a closer examination of publication patterns for the existing definitions suggest that this interest is not necessarily widespread within the field. Of the definitions provided for stimulability testing, the majority (71%) emerged from two author groups. Seven (41%) of the definitions were from articles articled by Gillespie and/or Gartner-Schmidt and colleagues,^6,12,20,21,31,32,50^ while five definitions (29%) were from articles authored by Toles and colleagues.^5,11,13,33^ The definitions provided by Gillespie, Gartner-Schmidt, and colleagues were more internally consistent than those provided by Toles and colleagues; indeed, four of the definitions from Gillespie, Gartner-Schmidt, and colleagues used essentially the same wording.^6,20,21,31^ Despite this, variability between definitions was still apparent, both within and, particularly, between author groups.

Thus, definitions of stimulability testing within the current peer-reviewed literature are limited in both number and authorship, yet also somewhat variable in content. These findings support the need for increased field-wide discussion of stimulability testing practices.

A secondary goal in collecting current definitions of stimulability testing was to determine the degree of consensus across definitions. While thematic analysis is not necessarily meant as a tool to determine group consensus, it is useful in that it can offer insight into recurring patterns within data (i.e., themes). Further, examining how frequently each theme appears across definitions can lend insight into the degree of consensus amongst authors regarding the need to include that theme within a definition of stimu ability testing.

To this end, thematic analysis of stimulability testing definitions resulted in five themes: Modifying Behaviors, Assessing Patient Response, Using Facilitators, Specifying Goals, and Contextualizing the Evaluation. Of the five themes that emerged, only the Modifying Behaviors theme appeared across all definitions, suggesting that the process of eliciting behavioral change from a patient during stimulability testing is, in the eyes of all authors, a universal requirement of a stimulability testing definition. The Assessing Patient Response (i.e., determining the patient’s ability to modify their behavior) and Using Facilitators (i.e., the use of models, cues, or other facilitating techniques) themes appeared in the majority (> 80%) of definitions, likewise suggesting a strong association between these elements and the concept of stimulability testing. The remaining themes, Specifying Goals and Contextualizing Evaluation, were present in the minority (≤35%) of definitions. While the reduced prevalence of these themes does not necessarily mean that they have reduced importance to defining stimulability testing, it does imply that they are not as universally considered when authors set forth to define stimulability testing in their work. Thus, the current results suggest most authors define stimulability testing using descriptions of modifying patient behaviors, assessing patient responses, and using facilitators.

Relative frequency is one method of determining the relative importance of themes when considering the “essential elements” of a stimulability testing definition. Another method is considering which themes serve to clearly differentiate stimulability testing from other aspects of a behavioral voice assessment. For instance, the Modifying Behaviors theme centers stimulability testing as a dynamic assessment, setting it apart from other static assessment procedures performed during voice evaluations (e.g., videostroboscopy). A definition of stimulability testing without this theme would not capture a core process of stimulability testing. Both the Assessing Patient Response and Using Facilitators themes similarly differentiate stimulability testing from other procedures. Assessing Patient Responses points to stimulability testing as an assessment procedure, (i.e., a procedure resulting in an outcome measure that will vary between individuals depending on their performance), which differentiates stimulability testing from treatment procedures which may share similar characteristics (i.e., stimulability testing using clear speech is not the same as Conversation Training Therapy). Using Facilitators implies the involvement of direct therapeutic intervention, rather than change occurring through indirect processes (e.g., hormonal changes, seasonal allergies, patient emotional state, etc.)

On the other hand, the remaining two themes (Specifying Goals and Contextualizing the Evaluation) do not currently have enough specificity to differentiate stimulability testing from other elements of a voice evaluation. For example, within the Specifying Goals theme, authors describe using stimulability testing for goals such as “guide treatment decisions”, “establish self-mastery” and “determine readiness for change”. While these goals for stimulability testing mirror those previously established in the literature,^3^ none are goals that can be uniquely achieved by stimulability testing or that differentiate stimulability testing from other aspects of a voice evaluation. One exception to this is the goal set out by Grillo (2021)^34^: “finding the best voice production techniques”^(p569)^. Since this goal was provided by only a single author, however, it is unclear the degree to which author authors and clinicians would agree this is the goal that sets stimulability testing apart from other elements of an evaluation. Similarly, the contexts provided within the Contextualizing the Evaluation theme were relatively few and extremely broad, ranging from contexts regarding timing of the evaluation (“performed during the pretreatment screening”^7^) to the timing of assessing the outcome measure (“ability to immediately change”^12^). Thus, this theme did not have enough internal cohesion to determine discriminating characteristics of stimulability testing.

Taken together, results of the thematic analysis suggest three “core” thematic elements for defining stimulability testing: modifying patient behaviors, assessing patient responses, and using facilitators. These thematic elements are supported not only by frequency of appearance in current peer-reviewed definitions of stimulability testing, but also because each conceptually differentiates stimulability testing from other behavioral assessment procedures. These thematic elements could therefore be used as a foundation for evaluating definitions of stimulability testing going forward; definitions that do not at least include descriptions of each element would not be aligned with the highest available standards for a stimulability testing definition. As research in this area continues, more stringent standards, perhaps surrounding specifying goals and contextualizing the evaluation, could be implemented to continue to establish what differentiates stimulability testing from other practices.

### Definitions of Stimulability Testing Outcomes

Seven articles (8%) provided a definition of stimulability testing outcomes; within these articles, ten different definitions were extracted. As with stimulability testing, most articles (57%) defining stimulability testing outcomes were published within the last 5 years, and all had been published within the last 15 years. Definitions of stimulability testing outcomes were also dominated by the same two authors groups; two definitions (20%) were provided by Gillespie, Gartner-Schmidt and colleages^12,20^ and five definitions (50%) were provided by Toles and colleagues.^5,11^ Also similar to the stimulability testing definitions, Gillespie, Gartner-Schmidt and colleagues provided relatively similar definitions across articles, while definitions provide by Toles and colleagues varied broadly. This again demonstrates the limited scope of publications in this area, supporting the need for increased dialogue and research on stimulability testing practices.

Thematic analysis was performed on the definitions of stimulability testing outcomes to aid in determining areas of consensus across definitions. However, unlike with stimulability testing, the results of the thematic analyses did not yield significant insights into commonalities across definitions. While two of the three themes identified (Defining Outcomes and Supporting Change) appeared across all ten definitions, the level of detail provided within definitions varied dramatically. For instance, some definitions described outcomes in an extremely broad manner (e.g., “change a behavior”)^14^, while others were more specific (“the degree of improvement perceived in the voice quality”).^11^ Further, sixteen unique outcomes were described across the ten definitions, only four of which were repeated across multiple definitions. Some articles described assessing only a single outcome measure, while other articles mentioned multiple outcomes. Similarly, some articles specified what types of support were given (e.g., “minimal cueing”)^51^ while others were more general (e.g., “models or cues”).^20^ Thus, while definitions shared thematic similarities, there was minimal agreement within each theme that would allow for statements regarding general consensus across definitions. In addition, the three themes generated using thematic analysis (Defining Outcomes, Quantifying Change, and Supporting Change) reflect expected aspects of defining measurements from psychometric and educational assessment perspectives.^52,53^ This suggests that the themes capture only basic elements of measurement science (i.e., what is the measurement and how is it measured), again suggesting broad thematic similarities without sufficient information for detailed consensus statements.

Overall, stimulability testing outcomes are rarely defined across the literature, and, when defined, definitions vary significantly from one another. While the current review did not find sufficient evident to suggest elements of consensus across definitions, the results of the thematic analysis do provide readers with a guide for effective definitions of stimulability testing outcomes going forward: including a clear definition of the outcome itself, descriptions of the context in which the outcome is measured and the rating scale with which the outcome is measured, and the type and degree of clinician support provided. Ensuring that each of these variables are clearly described when defining stimulability testing outcomes will improve transparency in stimulability testing reporting for both authors and clinicians, potentially allowing for identification of consensus elements of stimulability testing outcomes in future work.

### Behavioral Targets of Stimulability Testing

Behavioral targets were extractable in 54 (61%) of articles, including 35 articles (40%) that did not include formal definitions of stimulability testing or stimulability testing outcomes. These results imply that many authors consider it important to communicate the behavioral target of stimulability testing. This makes sense when considering the variety of behavioral targets displayed in Table 7 – after grouping similar targets together, a total of 31 different targets are listed across three categories (Behavior - general; Behavior - facilitator; and Individual). The most common listed target is improved voice quality (listed nine times across articles) followed by a three-way tie between forward focused resonance, increasing loudness, and voice therapy (each listed six times). Further, 20% of articles describe targeting across multiple classification types (e.g., determining both whether a patient was stimulable for “improved communication skills” as well as for “using intent”), and an additional 15% of articles specified multiple targets within a single classification type (e.g., targeting “forward focus” as well as “reduced tension of the neck/shoulders”^54^). The variety of targets seen within this review demonstrates that without language specifying the behavioral target(s) of stimulability testing, two SLPs performing stimulability testing could easily be performing radically different assessment tasks with extremely different clinical goals in mind.

As a form of dynamic assessment, stimulability testing can and should be tailored to the needs of an individual patient. Stimulability testing is valuable because it allows a clinician to weigh the needs of the patient against the potential facilitative tools at their disposal; a behavioral target that makes sense for one patient may not make sense for another. Thus, the range of behavioral targets presented in Table 7 is not inherently problematic. However, simply listing a target behavior for stimulability testing does not offer substantial detail regarding how changes in that behavior are measured, nor does it offer much insight into the clinician’s underlying conceptualization of stimulability testing. For example, stating that “stimulability for symptom improvement with voice optimization techniques”^55^ was performed gives some information regarding the target behavior (“symptom improvement”) and the context of assessing that behavior (“voice optimization techniques”). However, it does not give information regarding what constitutes “improvement” (e.g., changes in clinician-perceived vocal quality versus patient-perceived vocal pain or effort vs acoustic or aerodynamic metrics) nor a measurement scale used to quantify such changes (e.g., binary, categorical, continuous).

Further, it is unclear from this description whether the authors would consider patients with similar levels of improvement with different optimization techniques similarly stimulable, or whether there would be an additive effect of good performance on multiple facilitators. Thus, in many cases, the specification of a target behavior gives limited information regarding the outcome measures of stimulability testing, and how outcomes gathered by one author might differ conceptually from those gathered by another. Therefore, while specifying a behavioral target improves the overall specificity of stimulability testing descriptions, it is generally not sufficient to provide an accurate representation of an author’s conceptualization of stimulability testing.

### Measurement Scale Classification for Stimulability Testing Outcomes

Stimulability testing outcomes have been described using several different measurement scales, ranging from a binary scale (i.e., “stimulable” vs “not stimulable”) to continuous scales.

The results indicated that binary scales are the most used measurement scale type within the literature, occurring in 81% of articles, compared to 12% of articles using a categorical scale. This is the opposite trend compared to clinician reports; of the 88 practicing voice-specialized SLPs surveyed by Toles & Young (2023),^3^ only 23% reported using a binary scale, compared to 67% who reported using categorical scales. This highlights a divergence in how stimulability testing outcomes are described in research and clinical practice. As seen in the current review, there are multiple potential behavioral targets for stimulability testing; thus, for research purposes, it may simply be more straightforward to assign stimulability testing outcomes to the binary “stimulable” vs “not stimulable” classification. In clinical practice, however, where more nuanced decision-making is required based on a patient’s response, a categorical scale of measurement may be required. Regardless of the cause, divergences between published literature and clinical practice serve to underline areas where further investigation is needed.

### Terminology Used to Refer to Stimulability Testing and Stimulability Testing Outcomes

Readers may note that the definition of stimulability testing outcomes provided by Schneider (2019)^14^ in Table 5 appears more similar to definitions of stimulability testing, appearing in Table 3. Schneider’s^14^ definition was classified as one of stimulability testing *outcomes* rather than stimulability testing for two reasons. First, the wording within the definition implies that the term “stimulability”, is referring to an outcome measure (“stimulability is defined as *a patient’s ability to change a behavior”* [italics added]^14(p475)^). Modification of wording to read “stimulability is defined as *the assessment* of a patient’s ability to change a behavior” would imply that the term “stimulability” refers instead to stimulability testing. Second, the definition appeared immediately under the heading “Stimulability for Change”, implying that it was a definition for an outcome measure, rather than the assessment procedure. In contrast, definitions that used similar wording (e.g., see definition in Table 3 provided by Toles and Young, 2023^3^) but that were in contexts strongly suggesting an assessment procedure, were classified as definitions of stimulability testing. These cases do highlight the difficulty that the author encountered in many instances in differentiating whether terminology was used to refer to the testing procedure or the outcome measures. Throughout all articles, “stimulability testing” was the most frequently used terminology to refer to stimulability testing, while “stimulability” alone (i.e., without the use of further modifiers, such as “testing” or “probes”) was most the most common terminology to refer to stimulability testing outcomes. However, “stimulability” alone was used to refer to stimulability testing in 13% of articles, making it the second-most common term used to refer to stimulability testing. This likely points to an underlying conflation between the concepts of stimulability testing and stimulability testing outcomes, ultimately limiting the usefulness of the term “stimulability” for research and clinical practice purposes. There is therefore a need for clearer terminology usage when describing stimulability testing practices.

The author has used “stimulability testing” and “stimulability testing outcomes” throughout this review to refer to the assessment and assessment results of stimulability practices, respectively. This was done to maximize clarity when referring to these two separate concepts. However, the author acknowledges that “stimulability testing outcomes” is a somewhat cumbersome phrase. Alternate terminology that maintains the distinction between these two concepts is “stimulability testing” to refer to the procedure, and “stimulability level” to refer outcomes of the procedure. This terminology adds a modifier to the term “stimulability” in both cases, which both avoids the pitfalls of the “stimulability/stimulability testing” dichotomy outlined in the paragraph above. Further, by reinforcing the use of a modifier whenever using the term “stimulability”, authors and clinicians would be encouraged to considering which aspect of stimulability procedures they are describing. “Stimulability level” has successfully been used to differentiate stimulability testing and stimulability testing outcomes in previous work, suggesting potential for this term in work going forward.^5,11^ Regardless of which terminology is used, however, authors and clinicians should be mindful of clearly differentiating stimulability testing and stimulability testing outcomes within their work.

### Limitations

Limitations of this review must be acknowledged. First, the author opted to use the key terms “stimulability” and “stimulable” when searching for articles regarding this topic in the formal literature search. This was a deliberate choice to limit the scope of the current review to terminology that is known to be actively used and understood within current clinical practice.^3,4^ However, additional peer-reviewed literature exists examining practices similar to stimulability testing described using different terminology. For example, Dejonckere and Lebacq (2001)^22^ describe assessing “vocal plasticity”, and many subsequent authors cite this assessment practice when describing or defining stimulability testing.^3,6,12,13,32,56,57^ Without first establishing a formal definition of stimulability testing, however, it is unclear which of these differently named practices would be considered similar enough to stimulability testing to be included within the review. Future work could expand upon the current review by evaluating whether work describing practices similar to stimulability testing contained similar thematic elements to those described in the current review.

Further, the current review only included literature from peer-reviewed sources. This eliminated many potentially fruitful sources from consideration, including textbooks, theses, and dissertations. As noted in the introduction, many consider a textbook^2^ the basis of stimulability testing in voice disorders. Interestingly, however, Young (2025)^10^ found that descriptions of stimulability testing and stimulability testing outcomes from an informal review of voice textbooks were generally quite poor. This review examined 17 textbooks and found that only 71% of them (12/17) described a procedure resembling stimulability testing, and only 12% (2/17) described stimulability testing outcomes. Despite this, a formal review of addition of sources of information outside of peer-reviewed journal articles could be an interesting avenue for future work.

### Future Directions

The results of this literature review highlight many considerable gaps in the current literature regarding stimulability testing in voice disorders, suggesting the need for much future work. More than three quarters of the literature examined used terms such as “stimulability” or “stimulable” without providing a functional definition for either term. Definitions of stimulability testing and stimulability testing outcomes were available in less than a quarter (20%) of all examined literature; definitions of stimulability testing outcomes were particularly sparse. While many definitions shared thematic similarities, there was considerable variability in both the level and types of details provided between definitions. The current literature therefore provides enough consensus only to provide general guidelines for the definitions of stimulability testing and stimulability testing outcomes going forward; there is not currently sufficient consensus to support a standardized definition of either aspect of stimulability practices. Because of this, authors are called upon to ensure that future descriptions of stimulability testing, and particularly stimulability testing outcomes, are sufficiently detailed to support further development of standardized definitions.

Those that use stimulability testing regularly must also consider that the functions of stimulability testing, such as determining a patient’s candidacy for therapy, depend on a clinician’s ability to reliably and accurately assess the *outcomes* of stimulability testing.

Unfortunately, many of the reported functions of stimulability testing currently have extremely limited and frequently mixed literature supporting their validity in clinical practice.^5,12,13,22^ This may be in part to a failure to adequately describe stimulability testing outcomes within the literature. Four elements emerged from the thematic analysis of stimulability testing outcomes that could be described when considering these outcomes: the outcome itself, how change in the outcome is measured and contextualized, and how change in the outcome is supported.

The articles examined within this review most often described the outcome measure of interest and/or the measurement scale, but this information alone is not sufficient for outcomes to be fully replicable across studies. Further, outcome measures were rarely repeated across studies; in Table 7, 31 unique behavioral targets were identified across 54 articles, meaning a new outcome was likely measured in less than every 2 studies. Without adequate descriptions of stimulability testing outcomes, a single term (“stimulable”) can be used to describe a variety of conceptually different behavioral changes. Thus, the term itself loses much of its predictive value for aiding clinicians and researchers in establishing the relationship between stimulability testing outcomes and the functions of stimulability testing. There is therefore a great need for clinicians and researchers to mindfully and transparently report what is being measured during stimulability testing in the literature.

## Conclusion

The current review represents the first attempt to collect and evaluate the current literature on stimulability testing in the context of voice disorders. Stimulability testing has been used in clinical voice disorders since the 1970s, but descriptions of this practice began to appear within the peer-reviewed literature only within the last three decades. Publication on this topic has increased dramatically within the last decade; however, there remains a dearth of literature providing definitions of both stimulability testing and stimulability testing outcomes.

Thematic analysis of provided definitions of stimulability testing suggest that most authors describe the practice as involving modifying patient behaviors, assessing patient responses, and using facilitators. Definitions of stimulability testing outcomes shared thematic elements but differed greatly in the details provided within each theme. Behavioral targets of stimulability testing varied widely across the review, and included both individual-specific targets (e.g., “stimulable for therapy”) and behavior-specific targets (e.g., “stimulable for a better voice”; “stimulable for increased airflow”). Given the importance of adequately and validly quantifying stimulability testing outcomes to use it determine causative relationships between stimulability testing and many of the functions of stimulability testing, there is a great need for increased transparency in the literature when defining and describing stimulability testing and stimulability testing outcomes.

## Data Availability

All data produced in the present work are contained in the manuscript

## Acknowledgements

This project was partially funded through the University of Utah CSD Department Seed Grant for Innovative Research. Special thanks to Adrianna Shembel, who provided valuable feedback on early versions of this manuscript. The author has no conflicts of interest to disclose.

## Data Availability Statement

All data used for the creation of this article is available from the author upon reasonable request.

